# Sodium-Glucose Cotransporter 2 Inhibitors for Lithium-Associated Kidney Dysfunction in Mood Disorders: A Real-World Historical Cohort Study

**DOI:** 10.1101/2025.09.02.25334914

**Authors:** Mete Ercis, Idil Tarikogullari, Vanessa K. Pazdernik, Maria L. Gonzalez-Suarez, Raman Baweja, Alessandro Miola, Osama A. Abulseoud, Jonathan G. Leung, Susan L. McElroy, Alfredo B. Cuellar-Barboza, Michael J. Gitlin, Aysegul Ozerdem, Mark A. Frye, Balwinder Singh

## Abstract

**Introduction:** Sodium-glucose cotransporter-2 inhibitors (SGLT2i), initially developed for type 2 diabetes, have shown promise in improving renal outcomes in patients with and without diabetes. However, their effect on lithium-associated kidney dysfunction remains unknown.

**Methods:** This historical cohort study included patients from Mayo Clinic (2001-2023) with mood disorders who received lithium for ≥6 months and later used SGLT2i for ≥1 month. Data on SGLT2i use and lithium treatment were extracted from electronic health records. Serum creatinine values were used to calculate estimated glomerular filtration rate (eGFR) trajectories. Linear mixed-effects models with piecewise linear splines were used to estimate eGFR slopes before and after SGLT2i initiation, adjusted for age and sex.

**Results:** Fifty-six patients (mean age 57.4 years, 46.4% female), predominantly with bipolar disorder (86%), were included. The mean eGFR, measured nearest to SGLT2i initiation, was 77.9±26.0 mL/min/1.73 m^2^, and the mean duration of SGLT2i use was 19.5±17.8 months. Before SGLT2i initiation, eGFR declined at a rate of −1.43 mL/min/1.73 m^2^ per year (p<0.001). After initiation, eGFR increased by 0.69 mL/min/1.73 m^2^ per year, reflecting a +2.13 change (p=0.025). Sensitivity analyses, including only patients on lithium at SGLT2i initiation (n=22) or who had >1 year of SGLT2i use (n=29) showed similar, though non-significant, changes in slopes.

**Conclusion:** SGLT2i treatment was associated with a significant improvement in eGFR trajectory in patients with mood disorders who received long-term lithium therapy. These findings suggest a potential role for SGLT2is in mitigating lithium-associated kidney dysfunction and highlight the need for randomized controlled trials in this population.

## Introduction

Lithium is the first line treatment for bipolar disorder^1,2^ and an augmentation option for major depressive disorder (MDD).^3^ Despite its proven efficacy in preventing manic and depressive episodes ^2^ and reducing hospitalization,^4,5^ its long-term use is limited due to concern about kidney dysfunction.^6-8^ This risk is particularly evident in patients with concomitant risk factors for kidney dysfunction such as diabetes mellitus (DM) and hypertension.^9-13^ Balancing the benefits of mood stabilization against potential kidney damage presents a significant challenge,^14^ especially in patients with these comorbidities. Currently, there are no data on the optimal management of kidney dysfunction in patients with mood disorders treated with lithium.^9,15,16^ This present a clinical dilemma, as management is restricted to mitigating risk factor^11^ or discontinuing lithium.^9,15^

To date, no pharmacological interventions have been proven to slow lithium-associated kidney dysfunction.^16^ The effect of lithium discontinuation on long-term kidney function remains unclear, with some studies reporting improved estimated glomerular filtration rate (eGFR) slope,^17^ while others showed no significant change.^10^ The lack of effective treatment options for lithium-associated kidney disease highlights a critical unmet need in mental healthcare.^18^ This underscores the urgency for research into novel therapeutic approaches that can protect kidney function while preserving the benefits of lithium therapy.

Sodium-glucose cotransporter-2 inhibitors (SGLT2i), originally developed for type 2 diabetes mellitus (T2DM), lower blood glucose by inhibiting its reabsorption in the renal proximal tubules, thereby facilitating its excretion in the urine.^19^ Beyond glycemic control, SGLT2i have demonstrated efficacy in renoprotective properties in patients with chronic kidney disease (CKD), including with and without diabetes.^20-24^ SGLT2is reduce CKD progression and incidence of cardiovascular events in diverse populations, with consistent benefits observed across racial groups^25^ and different sexes^26^. While direct pathophysiological evidence is lacking, their effectiveness may be due to shared features with other forms of CKD, such as tubular injury and fibrosis.^27^ However, their efficacy in patients with mood disorders or those on long-term lithium therapy (LTLT) has not been studied.

Given the lack of treatment options for lithium-associated kidney dysfunction and the potential renoprotective effects of SGLT2is demonstrated in other populations, we aimed to investigate the impact of SGLT2i use on eGFR trajectory in patients with mood disorders who received LTLT. We hypothesized that SGLT2i use would be associated with an improved eGFR trajectory compared to the pre-treatment period.

## Patients and Methods

### Ethical Approvals and Consent

This study was approved by the Mayo Clinic Institutional Review Board (IRB#: 18-005449). Patients who provided Minnesota Research Authorization for the review of their health records for research purposes were included. Informed consent was not required for the passive use of health record review under these conditions for those who had provided research authorization.

### Study Population and Data Source

We identified adult patients (aged 18 years or older) diagnosed with mood disorders (based on ICD-9 and ICD-10 codes) who were prescribed an SGLT2i and had lithium exposure (determined by detectable serum lithium levels or prescription data) using the Mayo Data Explorer (MDE). The MDE is a web application that allows researchers to access and filter data from Mayo Clinic’s Unified Data Platform, which provides access to over 30 years of electronic health record (EHR) data, including diagnoses, medications, laboratory results, and clinical notes.

### Inclusion and Exclusion Criteria

Patients were eligible if they: (1) had a mood disorder diagnosis and received care at Mayo Clinic between January 1, 2001, and September 1, 2023; (2) were prescribed an SGLT2i (canagliflozin, dapagliflozin, empagliflozin, ertugliflozin); and (3) had lithium exposure. Patients were excluded if they had used lithium for less than 6 months, SGLT2is for less than 1 month, or lacked creatinine measurements both before and after SGLT2i initiation (Figure 1).

**Figure 1.**
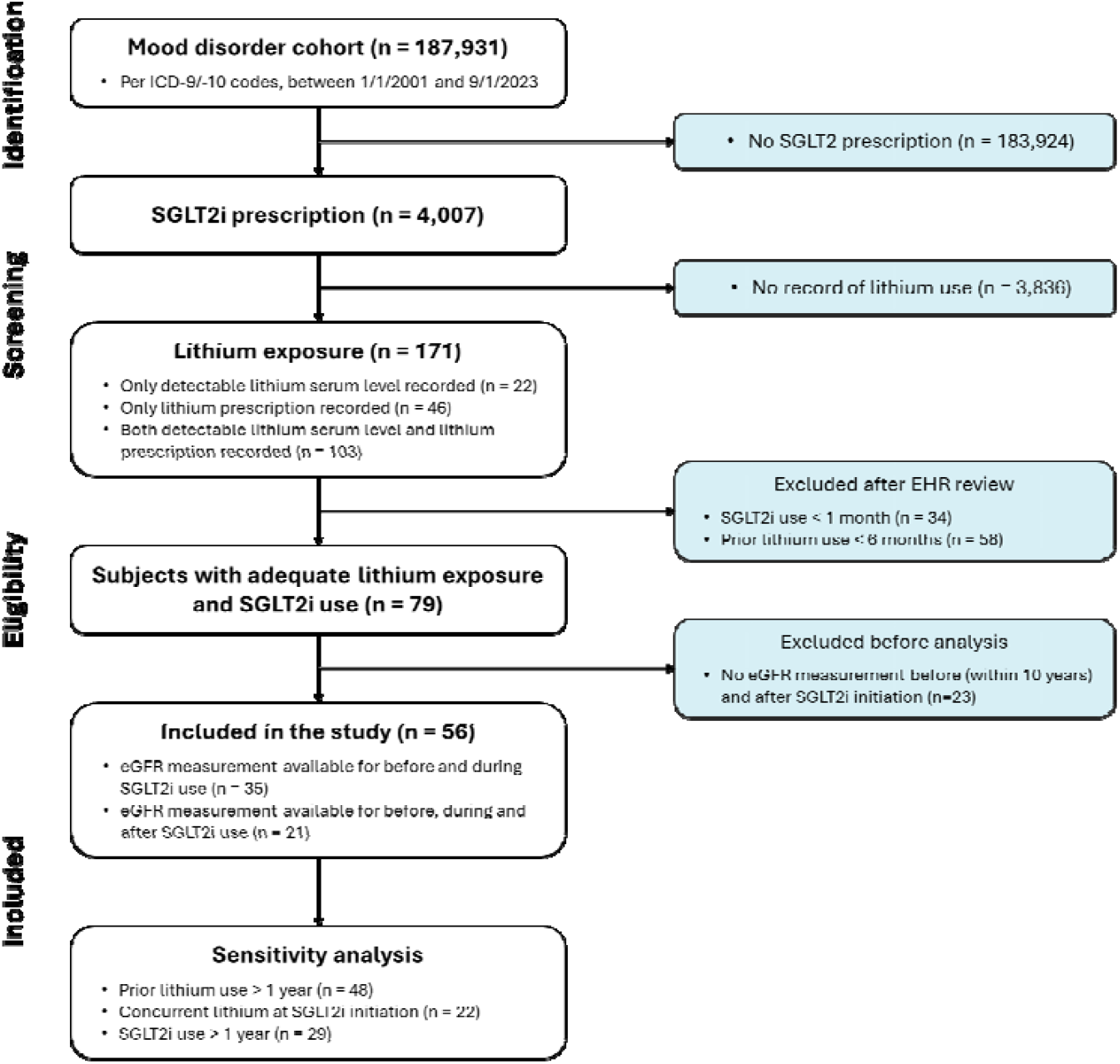
Flowchart of included patients. **Abbreviations:** eGFR, estimated glomerular filtration rate; ICD, International Classification of Diseases; SGLT2i, sodium-glucose co-transporter-2 inhibitor

### Data abstraction and eGFR calculation

Patient demographics (age, sex, race/ethnicity), clinical characteristics, dates of lithium and SGLT2i treatment, medical comorbidities, and concurrent medication use at the time of SGLT2i initiation were manually abstracted from EHR. Serum creatinine values were obtained using the MDE, and eGFR (mL/min/1.73 m^2^) was calculated using the 2021 CKD-EPI creatinine equation without the race coefficient.^28^ Only eGFR measurements recorded within 10 years prior to SGLT2i initiation and during SGLT2i treatment were included, excluding values after SGLT2i discontinuation.

### Outcomes

The primary outcome was the change in eGFR slope (mL/min/1.73 m^2^ per year) before and after SGLT2i initiation. This measure was chosen based evidence from clinical trials and observational studies demonstrating its utility in providing a meaningful estimate of kidney function and its ability to offer greater statistical power even in small populations with short follow-up periods.^29,30^

To assess the impact of SGLT2is on glycemic control in our sample, hemoglobin A1c (HbA1c) levels before and after SGLT2i initiation were also evaluated. The pre-SGLT2i HbA1c was selected from measurements taken within a window of 180 days before to 15 days after initiation (prioritizing values closest to the initiation date), while the post-SGLT2i HbA1c was selected from measurements within 75 to 180 days after initiation (prioritizing values closest to day 90 and excluding values obtained more than 15 days after discontinuation of the SGLT2i).

Among patients receiving concurrent lithium therapy, serum lithium levels before and after SGLT2i initiation were compared to evaluate the potential influence of SGLT2is on lithium clearance pharmacokinetics.

### Sensitivity analyses

Three sensitivity analyses were conducted to assess the robustness of the findings: (1) restricting the analysis to patients with >1 year of lithium use prior to SGLT2i initiation; (2) restricting the analysis to patients receiving concurrent lithium use at the time of SGLT2i initiation; and (3) restricting the analysis to patients with >1 year of SGLT2i use.

### Statistical analysis

Baseline characteristics of the cohort at the time of SGLT2i initiation were summarized using descriptive statistics. Continuous variables were reported as mean and standard deviation (SD) for normally distributed data and as range to describe the observed variation. For categorical variables, data were presented as absolute frequencies (n) and percentages (%). A multivariable linear regression model was used to assess demographic and lithium-related factors associated with eGFR at the time of SGLT2i initiation.

Linear mixed-effects models were used to examine changes in eGFR before and after SGLT2i initiation while adjusting for age and sex. Time was defined relative to SGLT2i initiation (time = 0), with a linear spline term allowing for a change in slope at treatment initiation. To account for within-subject correlation and individual variability, the model included random intercepts and random slopes for both time variables at the participant level.^31^ An exponential correlation structure was applied to model dependencies among repeated eGFR measurements within individuals.

Changes in HbA1c levels and serum lithium levels before and after SGLT2i initiation were assessed using the Wilcoxon signed-rank test with continuity correction.

All statistical analyses were performed using R Statistical Software (version 4.4.1; R Core Team 2022), with ‘*nephro’* package for calculating eGFR values from serum creatinine measurements and ‘*nlme*’ package for fitting linear mixed-effects models.

## Results

The study cohort comprised 56 patients (26 females, 46.4%, mean age 57.8 ± 11.9 years) with mood disorders (bipolar disorder, n = 48, 85.7%; and MDD, n = 7, 12.5%) (Table 1). The mean age at SGLT2i initiation was 57.8 ± 11.9 years and the majority of the patients identified as White (n = 54, 96.4%). The mean of the last measured eGFR prior to SGLT2i initiation was 77.82 ± 27.20 mL/min/1.73 m^2^, and 16 patients (28.5%) had stage 3 CKD based on this measurement. Most patients had a diagnosis of type 2 DM (n=50, 89.3%), which was the primary indication for SGLT2i use. The mean duration of lithium exposure prior to SGLT2i initiation was 7.04 ± 5.93 years, with 22 patients (39.3%) continuing lithium at SGLT2i initiation. The mean duration of SGLT2i use was 586.29 ± 533.50 days. A swimmer plot depicting individual timelines of SGLT2i and lithium use relative to initiation (time 0) is presented in Supplementary Figure 1. During the study period, 24 patients (42.9%) discontinued SGLT2i treatment. Among those who discontinued, the reason was not documented in 29.2% of cases. The most frequently documented reasons for discontinuation due to financial constraints (20.8%), lack of efficacy (12.5%), and genitourinary infections, comprised of urinary tract infections (16.7%) and genital mycotic infections (4.2%).

**Table 1.**
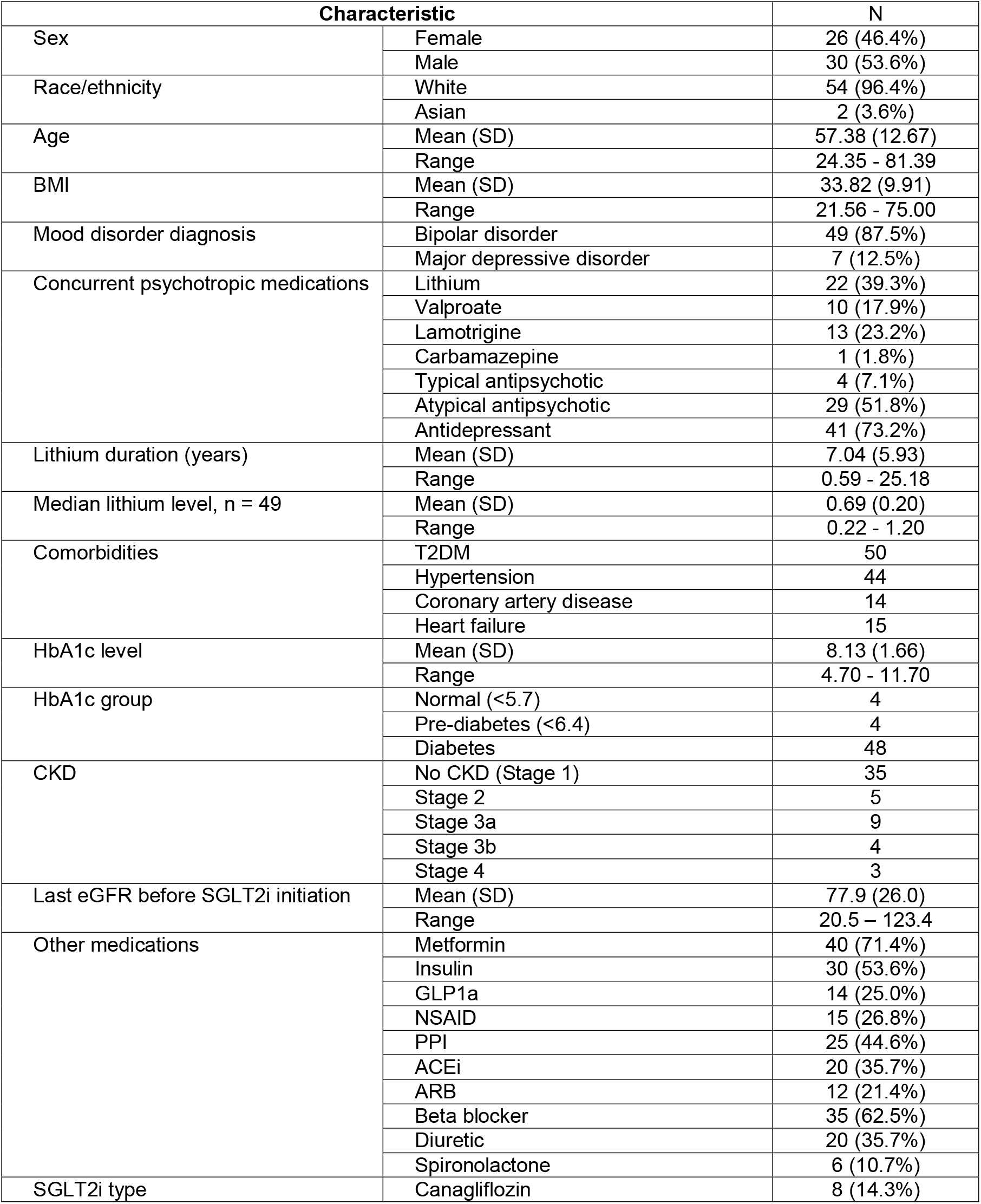
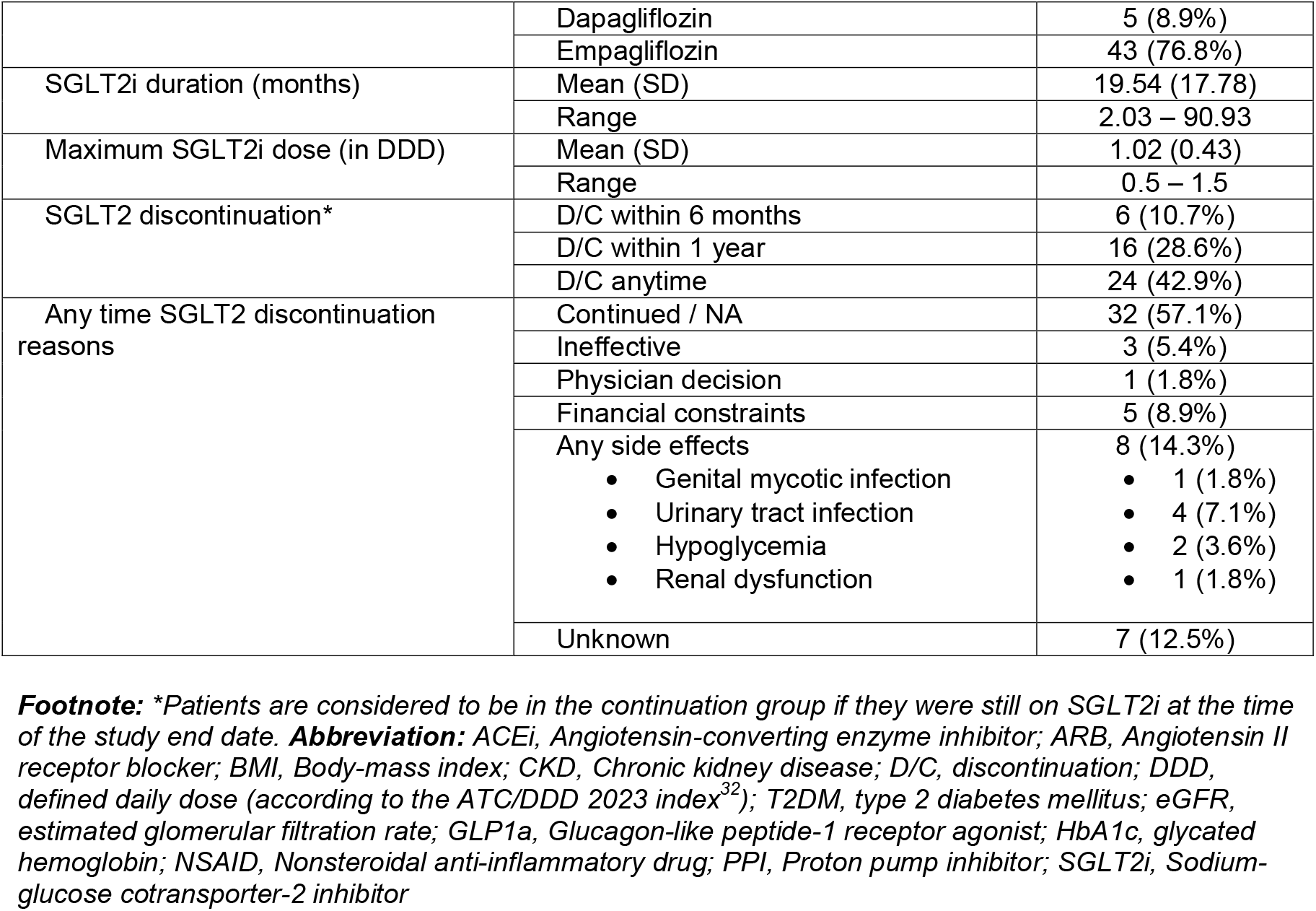
Patient demographics and disease characteristics at the time of SGLT2i start date (N=56)

Older age (β = −1.12, 95% CI: −1.62 to −0.61, p < 0.001) and higher median serum lithium levels (β = −36.59, 95% CI: −66.53 to −6.65, p = 0.018) were significantly associated with lower baseline eGFR. In contrast, sex (male: β = 8.00, 95% CI: −3.65 to 19.64, p = 0.173) and duration of lithium use (β = −0.72, 95% CI: −1.81 to 0.38, p = 0.194) were not significantly associated in this model.

### Primary outcome

A linear mixed-effects model with random intercepts and slopes, adjusting for sex and age, incorporated a linear spline for eGFR (knot at SGLT2i initiation) showed a significant decline in eGFR prior to SGLT2i initiation, with a mean slope of −1.43 (95% CI: −2.01 to −0.85, p < 0.001), indicating a 1.43-unit decrease in eGFR per year. After SGLT2i initiation, the eGFR trajectory improved significantly, with a change of +2.13 (95% CI: 0.27 to 3.98, p = 0.025) (Table 2, Figure 2). The post-SGLT2i initiation mean slope was +0.69 (95% CI: −0.90 to 2.28, p = 0.39). Older age at SGLT2i initiation was associated with a uniformly lower eGFR trajectory (β = −1.22, 95% CI: −1.59 to −0.84, p < 0.001), while sex was not a significant predictor (p = 0.732).

**Table 2.** eGFR slope estimates before and after SGLT2i initiation (N=56)

| ***Fixed Effects*** | ***Estimates*** | ***95% CI*** | ***p*** |
| --- | --- | --- | --- |
| *Intercept* | 147.99 | 125.65 – 170.33 | **<0.001** |
| eGFR slope (pre-SGLT2i) | -1.43 | -2.01 – -0.85 | **<0.001** |
| eGFR slope change (post-SGLT2i initiation) | 2.13 | 0.27 – 3.98 | **0.025** |
| Age at SGLT2i initiation | 1.22 | -1.59 – -0.84 | **<0.001** |
| Male | -1.63 | -11.12 – 7.86 | 0.732 |
| ***Random Effects (Variance Components)*** |  |  |  |
| *Intercept (SD)* | 20.7 |  |  |
| eGFR slope (pre-SGLT2i, SD) | 1.91 |  |  |
| eGFR slope change (post-SGLT2i initiation, SD) | 3.91 |  |  |
| Residual SD | 10.2 |  |  |
| ***Correlations*** |  |  |  |
| Intercept & eGFR slope (pre-SGLT2i) | 0.546 |  |  |
| Intercept & eGFR slope (post-SGLT2i initiation) | -0.228 |  |  |
| eGFR slope (pre- & post-SGLT2i initiation) | -0.776 |  |  |

**Abbreviations:** eGFR, estimated glomerular filtration rate; SGLT2i, sodium-glucose co-transporter-2 inhibitor

**Figure 2.**
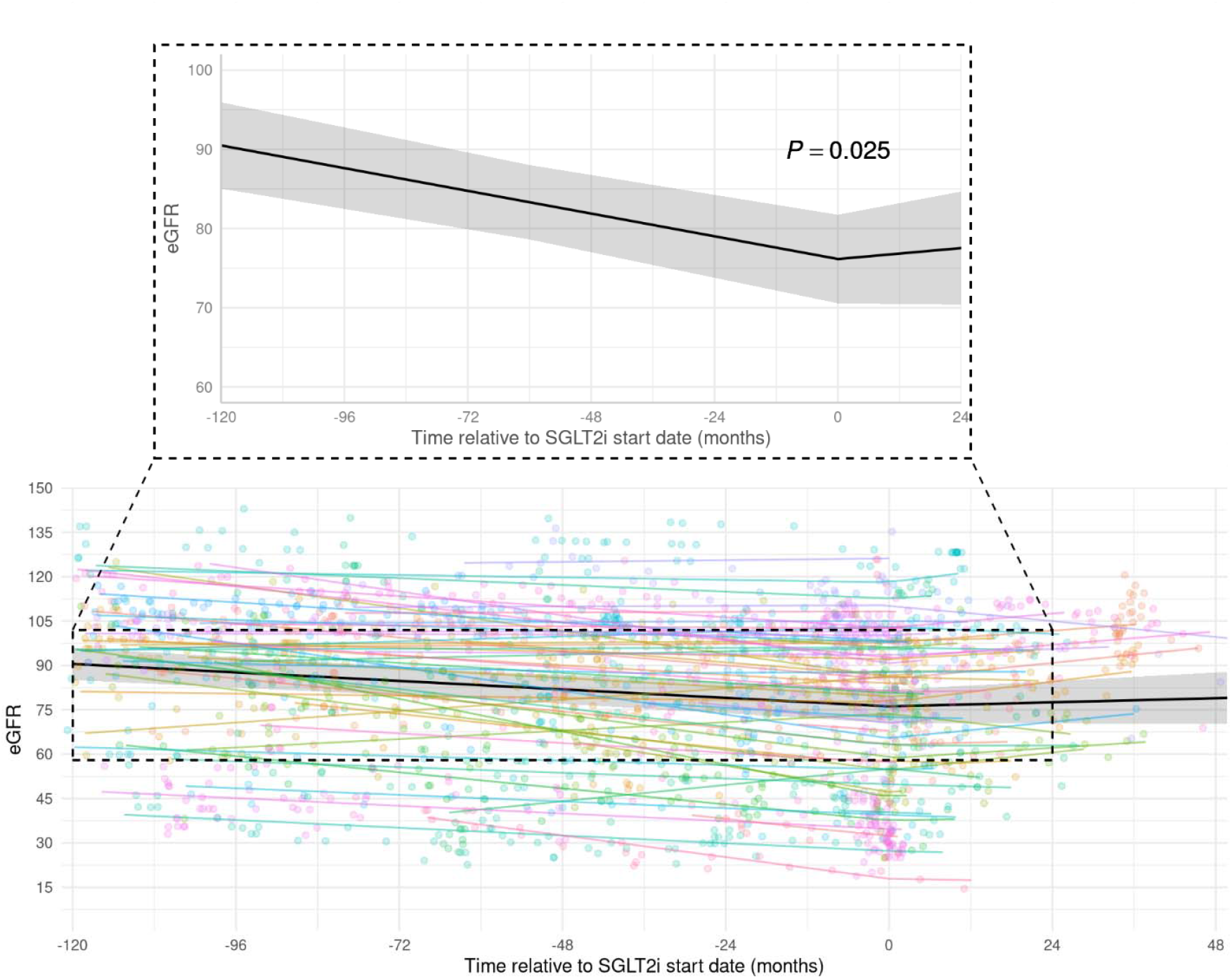
Observed and predicted eGFR trajectory before and after SGLT2i initiation. **Footnote:** Lower Panel: Individual patient eGFR measurements (colored dots) and trajectories (colored lines) are displayed in relation to the time of SGLT2i initiation (time = 0 months). The thick solid black line represents the overall model-predicted eGFR trajectory, derived from a linear mixed-effects model with a piecewise linear spline. Upper Panel: The model-predicted eGFR trajectory is highlighted, with the knot for the spline set at the time of SGLT2i initiation (time = 0 months), allowing for different slopes before and after this point. Shaded areas represent the 95% confidence intervals around the model-predicted trajectory.

### Secondary outcomes

Among patients with available pre- and post-SGLT2i HbA1c data (n = 24), the mean HbA1c decreased significantly from 8.5 ± 1.2% before initiation to 7.5 ± 1.1% after initiation, representing a mean reduction of 0.9 ± 1.5% (p = 0.009).

Serum lithium levels measured before and after SGLT2i initiation were available in a subset of patients (n = 11) receiving concurrent lithium therapy. The mean post-initiation level (0.70 ± 0.30 mEq/L) did not differ significantly from the mean pre-initiation level (0.66 ± 0.43 mEq/L, p = 0.721).

### Sensitivity analysis

Three sensitivity analyses were conducted to assess the robustness of the findings: (1) patients with >1 year of lithium use prior to SGLT2i initiation, (2) those receiving concurrent lithium therapy at the time of initiation, and (3) those with >1 year of SGLT2i use.

Restricting the analysis to patients with more than one year of lithium use prior to SGLT2i initiation (n = 48), the significant decline in eGFR before SGLT2i initiation persisted (β = −1.20, 95% CI: −1.82 to −0.57, p < 0.001). However, the improvement in eGFR trajectory after SGLT2i initiation, while similar in direction to the primary analysis, was not significant (β = 1.46, 95% CI: −0.37 to 3.29, p = 0.119) (Supplementary Table 1, Supplementary Figure 2).

When the analysis was restricted to patients receiving concurrent lithium therapy at the time of SGLT2i initiation (n = 22), neither the pre-SGLT2i eGFR decline (β = −0.82, 95% CI: −1.86 to 0.22, p = 0.122) nor the post-SGLT2i eGFR trajectory improvement (β = 1.21, 95% CI: −1.56 to 3.97, p = 0.391) was statistically significant, although both trends were consistent with the primary analysis (Supplementary Table 2, Supplementary Figure 3).

Lastly, in the sensitivity analysis restricted to patients with at least one year of SGLT2i use (n = 29), a significant eGFR decline before SGLT2i initiation was observed (β = −1.24, 95% CI: −2.10 to −0.38, p = 0.005). However, the subsequent change in eGFR trajectory after SGLT2i initiation was not statistically significant (β = 1.59, 95% CI: −0.56 to 3.75, p = 0.148) (Supplementary Table 3, Supplementary Figure 4).

Additionally, a test for interaction effect between concurrent lithium use at SGLT2i initiation and change in eGFR slope was not significant (p = 0.89), suggesting that the impact of SGLT2i on eGFR trajectory did not differ based on lithium status at initiation (Supplementary Table 4, Supplementary Figure 5).

## Discussion

In this historical cohort study, we observed that SGLT2i use was associated with a significant improvement in eGFR trajectory of patients with mood disorder on LTLT in this population. This novel finding provides the first evidence that SGLT2is may represent a promising option to mitigate lithium-associated kidney dysfunction, a condition with limited treatments and significant clinical challenges. Our findings are particularly relevant given the limited treatment options for lithium-associated kidney disease and the risks associated with lithium discontinuation,^9^ including mood episode relapse^5,12,33,34^ and increased suicide risk.^35^

Currently, the management of lithium-associated kidney disease is limited and the evidence regarding the benefit of lithium discontinuation is conflicting.^12,16,30^ For example, one population-based historical cohort study reported that in patients with mood disorders without pre-existing CKD, kidney function returned to baseline after lithium discontinuation.^12^ However, in patients who *had* already developed CKD, discontinuing lithium did not lead to significant improvement; rather, progressive renal function decline persisted despite cessation.^12^ In contrast, a mirror-image study using data from the LiSIE retrospective cohort found that stopping lithium slowed mean eGFR decline, with a difference attributable to discontinuation of +1.55 mL/min/1.73 m^2^/year (95% CI: 1.23 –1.87, p < 0.001).^30^ This effect was reportedly more pronounced in participants with lower baseline eGFR.^30^ These conflicting findings highlight the lack of certainty about the impact of discontinuing lithium on long-term renal outcomes. Although our findings should be interpreted cautiously due to the relatively small sample size and comorbidity profile (given nature of FDA-approved indications for SGLT2is), the observed eGFR slope improvement of +2.13 mL/min/1.73 m^2^/year (p = 0.025) after SGLT2i initiation, suggest a promising strategy to preserve kidney function while maintaining mood stabilization.

While SGLT2is have demonstrated renoprotective effects across adult populations with type 2 DM, heart failure, or CKD,^24^ there is currently no evidence regarding their effectiveness in patients with mood disorders—particularly those receiving LTLT. To date, no studies have specifically investigated their use in patients with mood disorders who had LTLT. The significant eGFR slope improvement observed after SGLT2i initiation in our real-world lithium-treated cohort is directionally consistent with the +0.23 mL/min/1.73 m^2^/year (95% CI, 0.05 – 0.40) reported for the EMPA-kidney trial’s chronic maintenance phase with empagliflozin.^36^ Our findings suggest that the renoprotective effects of SGLT2is may extend to patients with mood disorders undergoing LTLT, offering a potential therapeutic option for this population with an unmet need. Furthermore, our sensitivity analysis, restricted to patients receiving concurrent lithium therapy at the time of SGLT2i initiation, showed a similar trend toward eGFR improvement. This finding suggests that the renoprotective effects of SGLT2is observed in the overall cohort may also apply to patients who continue lithium therapy. This is particularly relevant for patients who respond well to lithium but are at risk of lithium-associated kidney dysfunction, as it may provide a strategy to continue this essential mood-stabilizing treatment while potentially preserving kidney function.

A key consideration is the potential for pharmacokinetic interactions between SGLT2is and lithium. The direct effects of SGLT2is on lithium clearance remain unknown, although some evidence suggests empagliflozin-mediated increases in lithium excretion.^37^ Given lithium’s narrow therapeutic index, these alterations could have significant clinical consequences, as subtherapeutic lithium levels may increase the risk of mood episode relapse. Although we did not identify a significant difference in patients’ serum lithium levels, our sample may not have been sufficiently powered to detect such changes.

Patients with bipolar disorder often experience a high cardiometabolic burden, including elevated rates of DM and hypertension. The positive findings from our real-world study are promising, suggesting that SGLT2is may offer a potential treatment option for patients with mood disorders receiving LTLT who also have comorbid diabetes and hypertension— addressing a common clinical dilemma faced by clinicians. Future randomized clinical trials including individuals without such comorbidities and with an established diagnosis of lithium-associated kidney dysfunction will be essential in establishing the causal relationship between SGLT2i use and the improvement of kidney function and determining the optimal dose and duration of SGLT2i treatment. Furthermore, the long-term efficacy and safety of SGLT2i use in patients on lithium need to be carefully evaluated.

It is also important to note that the anytime discontinuation rate for SGLT2i was relatively high at 42.9%. Although this rate is higher than those reported in clinical trials,^20,22,36^ it is comparable to the real-world at least one discontinuation rates (ranging from 37.7 to 56%).^38,39^ Similarly, the 6-month discontinuation rate in our cohort was 17.9%, which aligns with the literature (ranging from 6.1% to 20.1% at 6 months).^38,40^ The most commonly documented reasons for discontinuation were genitourinary infections and financial constraints. Future studies are necessary to better estimate the risk of certain side effects, such as genitourinary infections, in this population. These risks might be elevated due to high type 2 DM rates in our cohort. In addition, financial constraints and high out-of-pocket cost of SGLT2is have been identified as important reasons for discontinuation in other real-world studies,^38,41,42^ raising concerns about healthcare access, which may be especially critical for patients with severe mental illnesses such as bipolar disorder.^43,44^

### Strengths and Limitations

Our study’s strengths include its novel focus and the use of linear mixed-effects models to assess the entire eGFR trajectory for each patient. However, several limitations exist. First, due to its retrospective, observational design and lack of a control group, causal inference is limited. Second, this study was conducted at a single academic center with a predominantly White population, limiting its generalizability to non-White populations. While prior studies suggested SGLT2is provide similar benefits across racial groups,^25^ more research is needed. Third, the relatively small sample size may have reduced statistical power, particularly in sensitivity analyses, potentially limiting our ability to detect significant differences in eGFR trajectory in specific subgroups and also precluding subgroup analysis for patients with stage 3 or greater CKD. Fourth, the absence of a control group limits our ability to isolate the specific effect of SGLT2is. While matching on all potential confounders would be ideal, this was not feasible given the complex comorbidities and varying indications for SGLT2i use in this population. Future studies employing a negative control group (e.g., patients receiving other antidiabetic agents) could help clarify the specific association between SGLT2i use and kidney function improvements.

## Conclusion

In this real-world cohort study, we provide the first evidence that initiation of SGLT2i is associated with a significant improvement in eGFR trajectory among patients with mood disorders who received/receiving LTLT. These findings suggest SGLT2is may help mitigate lithium-associated kidney dysfunction—a condition for which no established treatments currently exist. Further randomized trials are warranted to confirm these findings, determine the optimal dosing and duration of SGLT2i therapy, and evaluate long-term efficacy, safety, and potential impact on mood symptoms in this population.

## Data Availability

The data that support the findings of this study are available on request from the corresponding author. The data are not publicly available due to privacy or ethical restrictions.

## Acknowledgements

This work was supported by the Department of Psychiatry and Psychology, Mayo Clinic, Rochester, Minnesota, USA. This publication was made possible by the Mayo Clinic CTSA through grant number UL1TR002377 and KL2 TR00237 from the National Center for Advancing Translational Sciences (NCATS), a component of the National Institutes of Health (NIH). Its contents are solely the responsibility of the authors and do not necessarily represent the official views of the NIH. The funding sources had no role in the design of the study, data collection, analysis or interpretation, writing of the manuscript, or the decision to submit the manuscript for publication.

