## Supplement file for "Sodium-Glucose Cotransporter 2 Inhibitors for Lithium-Associated Kidney Dysfunction in Mood Disorders: A Real-World Historical Cohort Study"

Supplementary Material Content List

### **Supplementary Tables**

Supplementary Table 1. eGFR slope estimates before and after SGLT2i initiation in patients with more than one-year lithium exposure prior to SGLT2i initiation (n = 48)

| ***Fixed Effects*** | *Estimates* | *CI* | *p* |
| --- | --- | --- | --- |
| *Intercept* | 145.95 | 119.62 – 172.27 | **<0.001** |
| eGFR slope (pre-SGLT2i) | -1.2 | -1.82 – -0.57 | **<0.001** |
| eGFR slope change (post-SGLT2i initiation) | 1.46 | -0.37 – 3.29 | 0.119 |
| Age at SGLT2i initiation | -1.15 | -1.59 – -0.71 | **<0.001** |
| Male | -2.51 | -13.11 – 8.10 | 0.636 |
| **Random Effects** |  |  |  |
| *Intercept (SD)* | 21.624 |  |  |
| eGFR slope (pre-SGLT2i, SD) | 1.938 |  |  |
| eGFR slope change (post-SGLT2i initiation, SD) | 3.999 |  |  |
| Residual SD | 9.285 |  |  |
| **Correlations** |  |  |  |
| Intercept & eGFR slope (pre-SGLT2i) | 0.554 |  |  |
| Intercept & eGFR slope (post-SGLT2i initiation) | -0.268 |  |  |
| eGFR slope (pre- & post-SGLT2i initiation) | -0.839 |  |  |

Supplementary Table 2. eGFR slope estimates before and after SGLT2i initiation in patients receiving concurrent lithium therapy at the time of SGLT2i initiation (n = 22)

| ***Fixed Effects*** | *Estimates* | *CI* | *p* |
| --- | --- | --- | --- |
| *Intercept* | 148.17 | 110.63 – 185.71 | **<0.001** |
| eGFR slope (pre-SGLT2i) | -0.82 | -1.86 – 0.22 | 0.122 |
| eGFR slope change (post-SGLT2i initiation) | 1.21 | -1.56 – 3.97 | 0.391 |
| Age at SGLT2i initiation | -1.06 | -1.71 – -0.41 | **0.003** |
| Male | -15.98 | -31.79 – -0.17 | **0.048** |
| **Random Effects** |  |  |  |
| *Intercept (SD)* | 19.689 |  |  |
| eGFR slope (pre-SGLT2i, SD) | 2.172 |  |  |
| eGFR slope change (post-SGLT2i initiation, SD) | 4.213 |  |  |
| Residual SD | 9.416 |  |  |
| **Correlations** |  |  |  |
| Intercept & eGFR slope (pre-SGLT2i) | 0.437 |  |  |
| Intercept & eGFR slope (post-SGLT2i initiation) | -0.139 |  |  |
| eGFR slope (pre- & post-SGLT2i initiation) | -0.929 |  |  |

Supplementary Table 3. eGFR slope estimates before and after SGLT2i initiation in patients with at least one-year SGLT2i use (n=29)

| ***Fixed Effects*** | *Estimates* | *CI* | *p* |
| --- | --- | --- | --- |
| *Intercept* | 147.09 | 109.27 – 184.91 | **<0.001** |
| eGFR slope (pre-SGLT2i) | -1.24 | -2.10 – -0.38 | **0.005** |
| eGFR slope change (post-SGLT2i initiation) | 1.59 | -0.56 – 3.75 | 0.148 |
| Age at SGLT2i initiation | -1.16 | -1.83 – -0.49 | **0.002** |
| Male | -5.66 | -19.90 – 8.59 | 0.422 |
| **Random Effects** |  |  |  |
| *Intercept (SD)* | 22.747 |  |  |
| eGFR slope (pre-SGLT2i, SD) | 2.174 |  |  |
| eGFR slope change (post-SGLT2i initiation, SD) | 4.478 |  |  |
| Residual SD | 9.357 |  |  |
| **Correlations** |  |  |  |
| Intercept & eGFR slope (pre-SGLT2i) | 0.554 |  |  |
| Intercept & eGFR slope (post-SGLT2i initiation) | -0.251 |  |  |
| eGFR slope (pre- & post-SGLT2i initiation) | -0.875 |  |  |

Supplementary Table 4. eGFR slope estimates before and after SGLT2i initiation with interaction effect of concurrent lithium therapy (n=56)

| ***Fixed Effects*** | *Estimates* | *CI* | *p* |
| --- | --- | --- | --- |
| *Intercept* | 148.89 | 126.03 – 171.74 | **<0.001** |
| eGFR slope (pre-SGLT2i) | -1.43 | -2.01 – -0.86 | **<0.001** |
| eGFR slope change (post-SGLT2i initiation) | 2.02 | -0.22 – 4.25 | 0.077 |
| Lithium at index date | -2.74 | -12.62 – 7.14 | 0.580 |
| Age at SGLT2i initiation | -1.22 | -1.59 – -0.84 | **<0.001** |
| Male | -1.59 | -11.15 – 7.97 | 0.740 |
| eGFR slope change (post-SGLT2i initiation) * Lithium at index date | 0.24 | -2.99 – 3.46 | 0.886 |
| **Random Effects** |  |  |  |
| *Intercept (SD)* | 20.904 |  |  |
| eGFR slope (pre-SGLT2i, SD) | 1.911 |  |  |
| eGFR slope change (post-SGLT2i initiation, SD) | 3.958 |  |  |
| Residual SD | 10.237 |  |  |
| **Correlations** |  |  |  |
| Intercept & eGFR slope (pre-SGLT2i) | 0.553 |  |  |
| Intercept & eGFR slope (post-SGLT2i initiation) | -0.221 |  |  |
| eGFR slope (pre- & post-SGLT2i initiation) | -0.75 |  |  |

### **Supplementary Figures**

Supplementary Figure 1. Swimmer plot showing SGLT2i treatment and lithium exposure relative to SGLT2i initiation (time 0) for each participant (n = 56). Colors indicate treatment status: lithium (blue), none (yellow), SGLT2i only (red), and lithium + SGLT2i (grey). For simplicity, any intermittent lithium discontinuations between the initial lithium start and final end dates are not shown and are instead presented as continuous treatment. Black dots indicate the first and last available eGFR measurements.

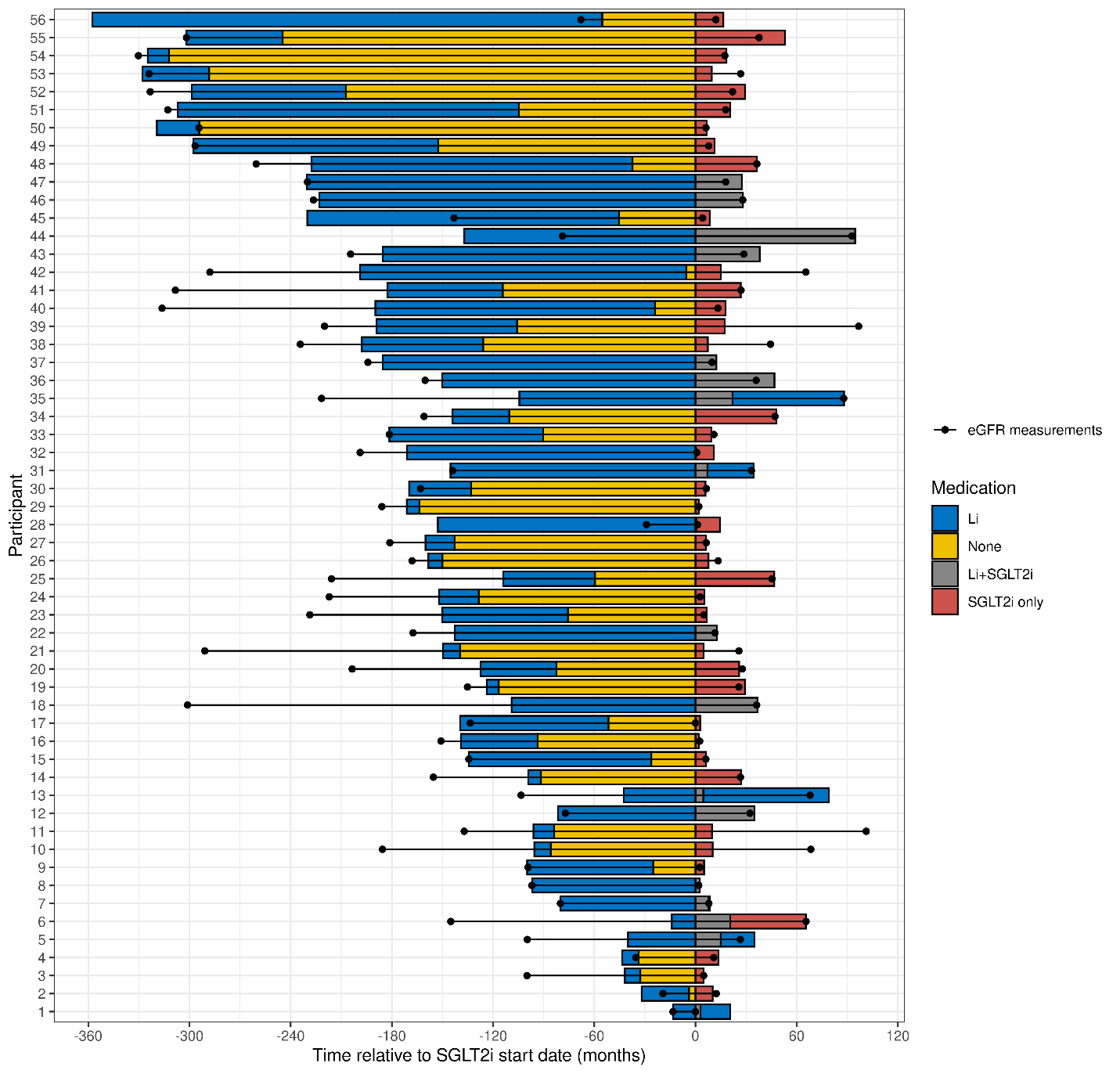

Supplementary Figure 2. Observed and predicted eGFR trajectory before and after SGLT2i initiation in patients with more than one-year lithium exposure prior to SGLT2i initiation (n = 48)

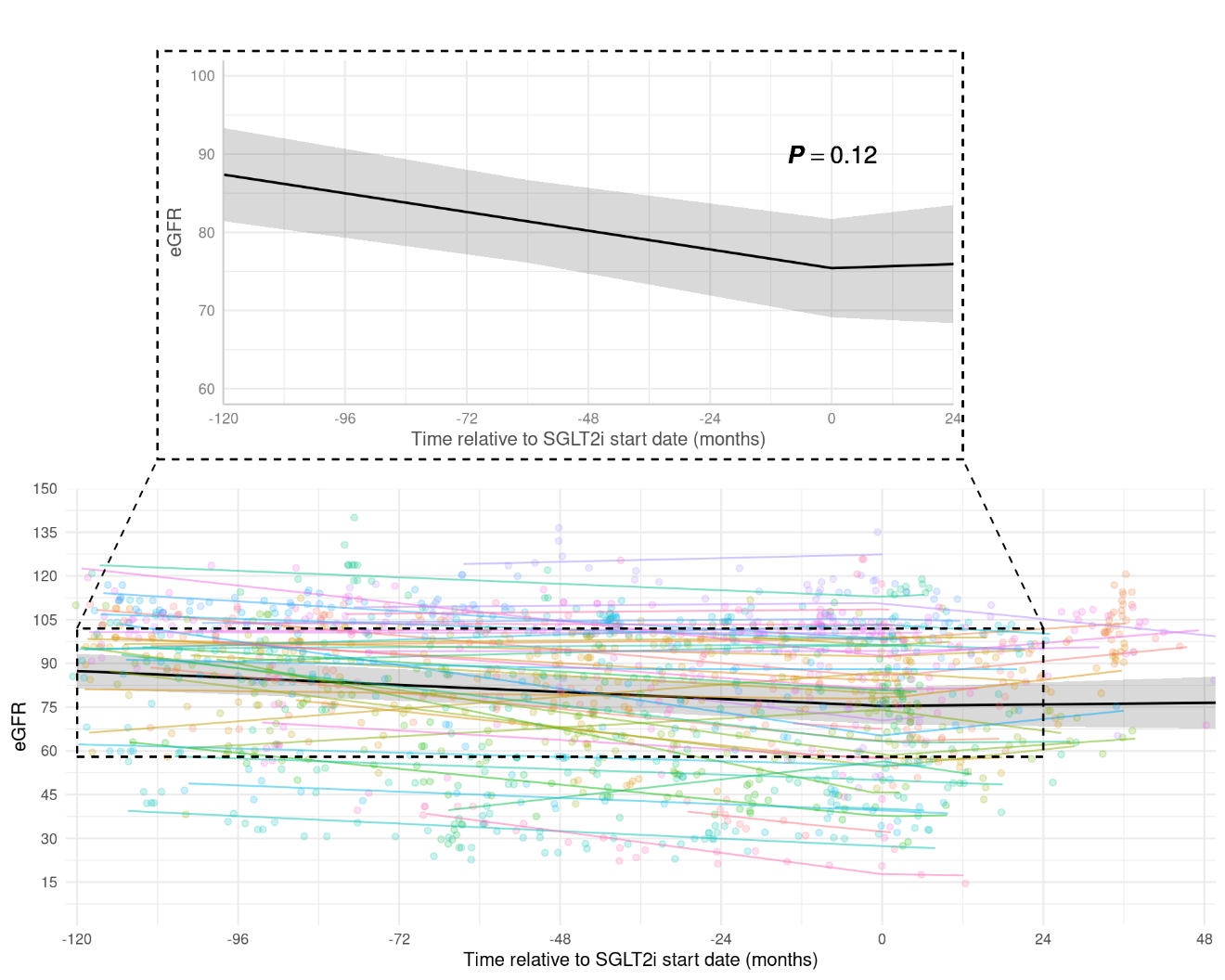

Supplementary Figure 3. Observed and predicted eGFR trajectory before and after SGLT2i initiation in patients receiving concurrent lithium therapy at the time of SGLT2i initiation (n = 22)

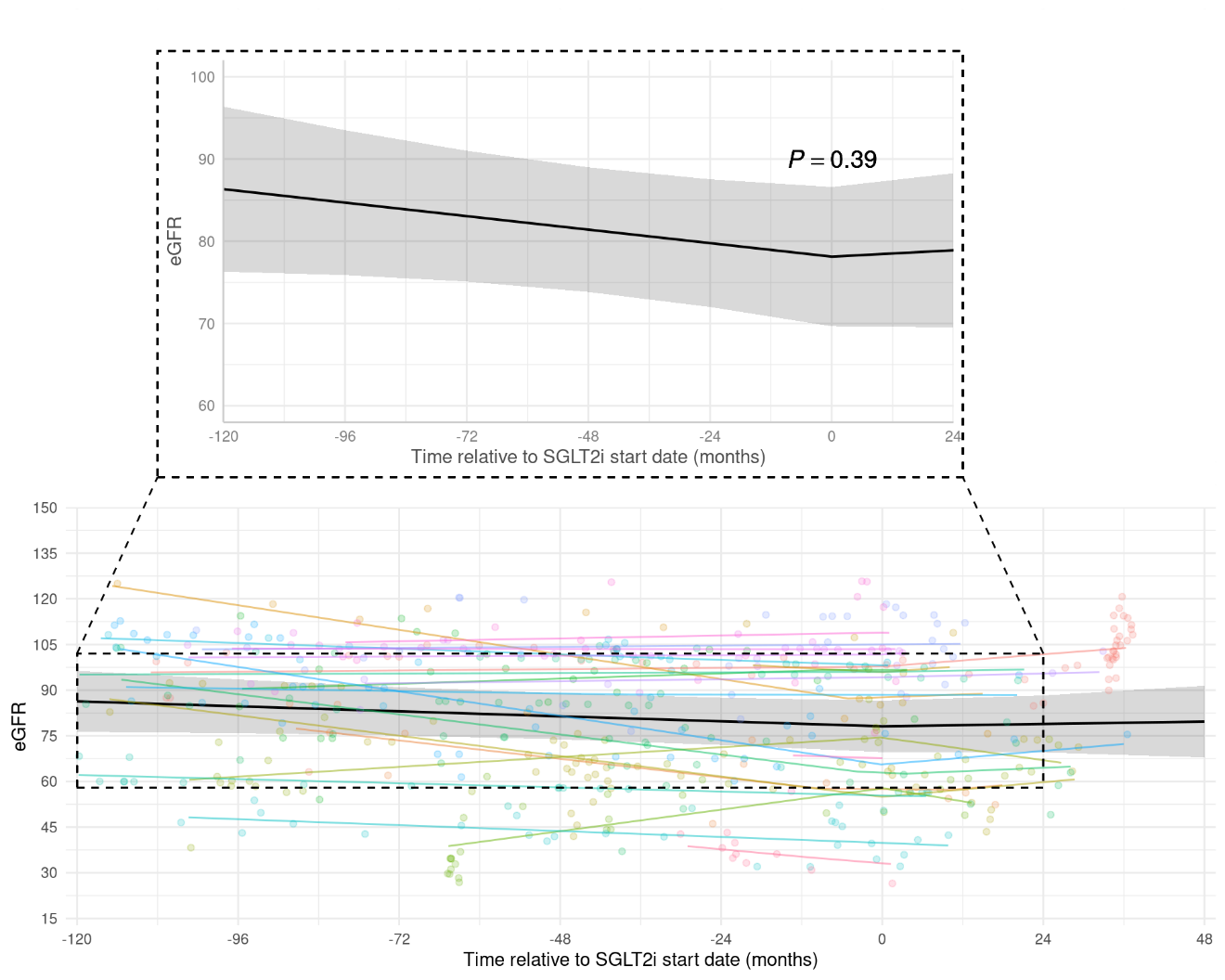

Supplementary Figure 4. Observed and predicted eGFR trajectory before and after SGLT2i initiation in patients with at least one-year SGLT2i use (n = 29)

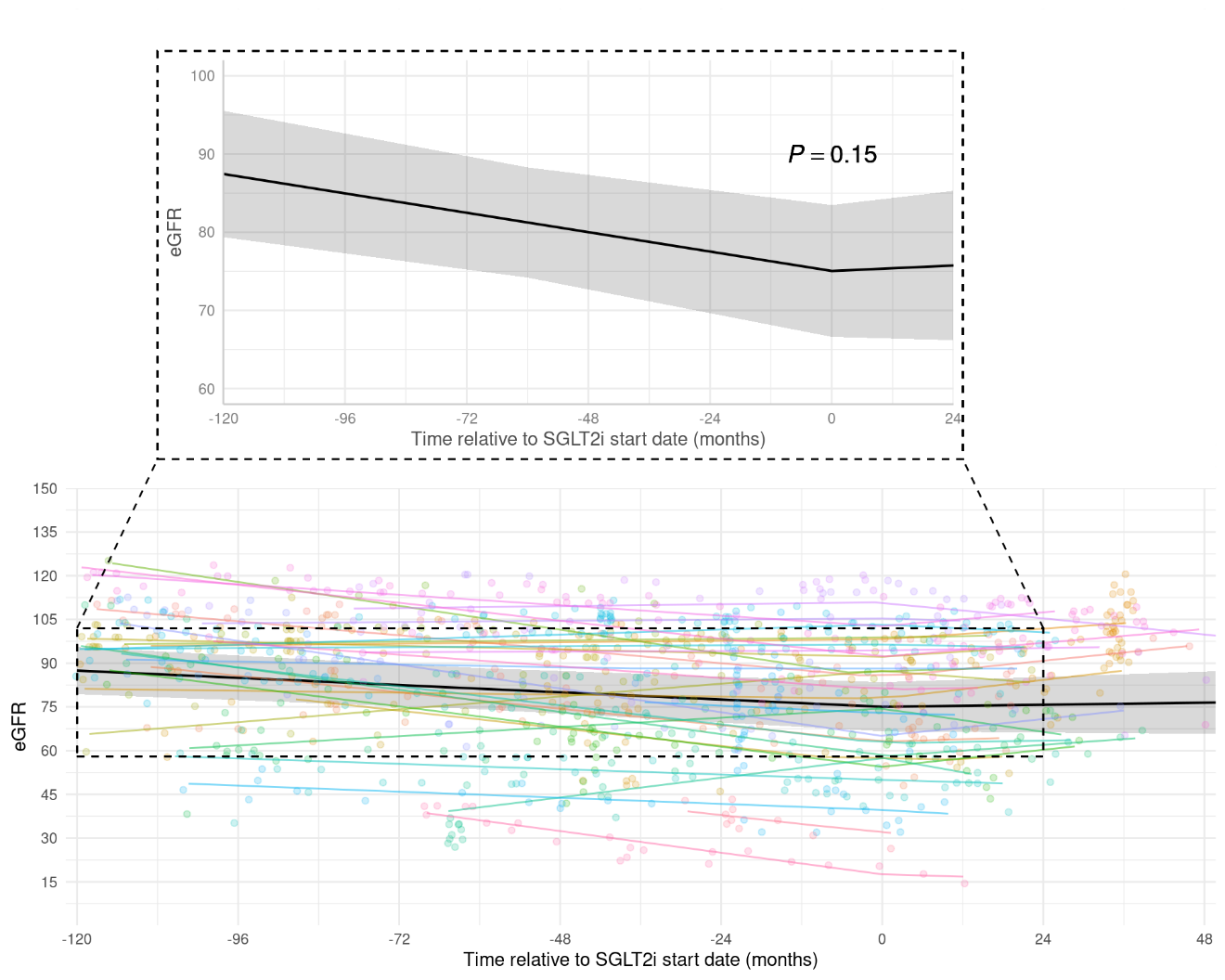

Supplementary Figure 5. Observed and predicted eGFR trajectory before and after SGLT2i initiation with interaction effect of concurrent lithium therapy (n=56)

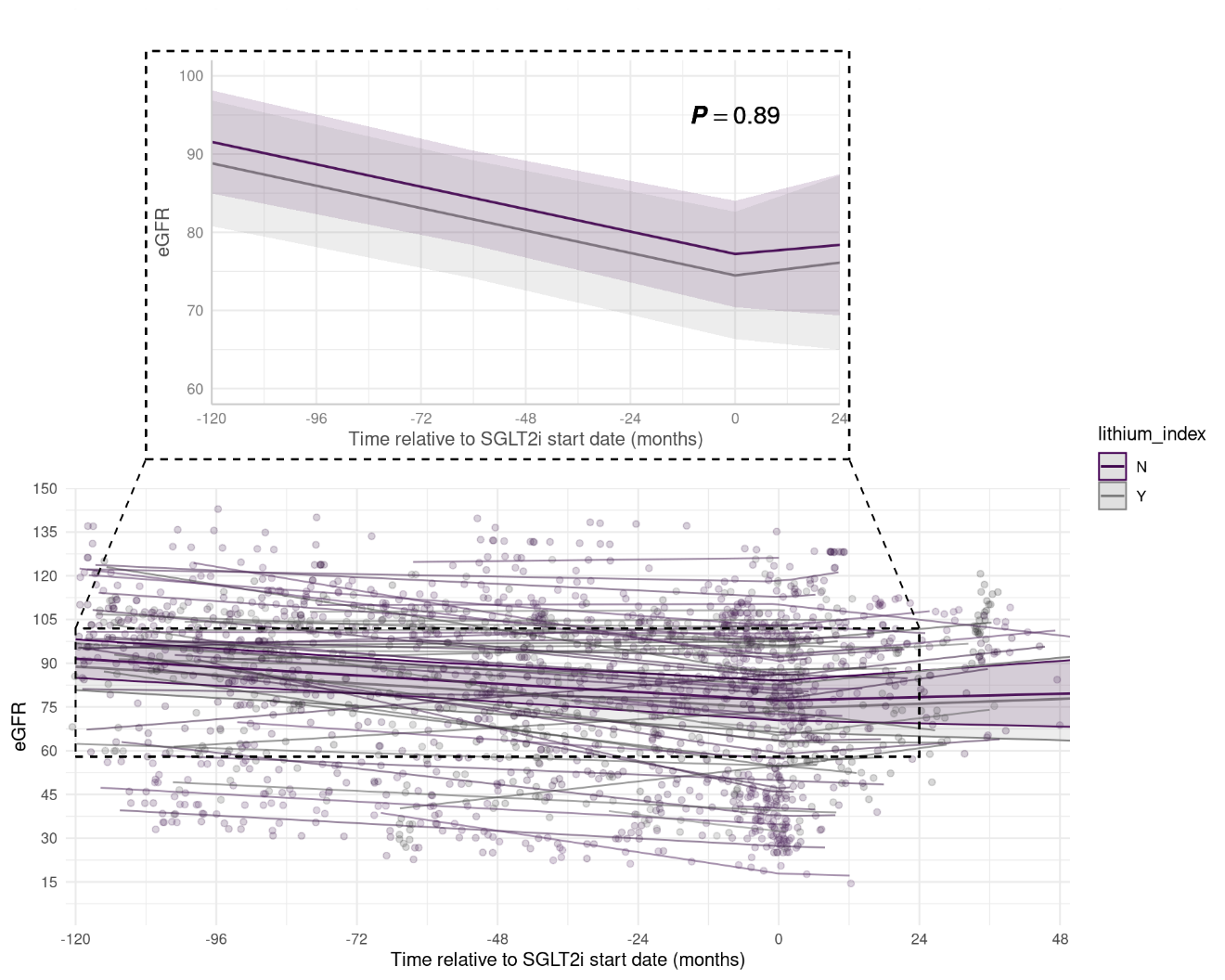
